# Incidental Non-Breast Malignancies in a Consecutive Forensic Autopsy Cohort: Secondary Findings from the Sisyphus Study

**DOI:** 10.64898/2026.05.05.26352437

**Authors:** Zacharoula Sidiropoulou, Carlos Santos

## Abstract

**Background:** Forensic autopsy cohorts can help estimate the burden of clinically unrecognised cancer that is not captured by routine incidence statistics. We report incidental non-breast malignancies identified as secondary findings in the Sisyphus Study, a prospective forensic autopsy cohort originally established to investigate silent breast cancer prevalence.

**Methods:** This was a descriptive secondary analysis of 291 consecutive medicolegal autopsies performed in Lisbon, Portugal, between July 2016 and December 2019 (74 male and 217 female decedents). Key exclusions relevant to the present analysis were age below 40 years, major breast-region injury, and known or clinically evident cancer. An incidental cancer was defined as a histologically confirmed malignancy identified at autopsy in an individual without a prior clinical cancer diagnosis.

**Results:** Fifteen incidental non-breast malignancies were identified among 291 decedents, yielding an overall prevalence of 5.15%. Prevalence was 6.76% in males (5/74) and 4.61% in females (10/217). Male findings comprised two colorectal adenocarcinomas, one pancreatic metastatic adenocarcinoma, one gastric adenocarcinoma, and one splenic lymphoma. Female findings comprised six colorectal adenocarcinomas, two lung adenocarcinomas, one perforated gastric adenocarcinoma, and one ovarian metastatic adenocarcinoma. Colorectal malignancies accounted for 8 of 15 cases (53.3%). Metastatic disease was documented in at least five cases, and perforation was present in two gastrointestinal tumours. None of the affected individuals had a prior cancer diagnosis during life.

**Conclusions:** This cohort demonstrates a measurable burden of clinically silent non-breast cancer, including advanced and potentially fatal disease. Forensic autopsy surveillance may complement conventional cancer surveillance by identifying malignancies that remain invisible to clinical registries. The predominance of colorectal cancer in this series is consistent with missed opportunities for earlier detection, although individual screening histories were unavailable.

## Introduction

Cancer surveillance is fundamentally based on clinically recognised disease. By design, registry-derived incidence counts exclude malignancies that remain undiagnosed during life. Autopsy-based research therefore offers a distinct view of the underlying reservoir of occult cancer, particularly when sampling occurs outside highly selected hospital populations (1,2).

Most autopsy literature on clinically silent cancer has focused on breast pathology. Classic medicolegal and autopsy studies, together with a later systematic review and meta-analysis, showed that post-mortem examination can identify a reservoir of otherwise unrecognised breast neoplasia and precursor lesions that is not apparent from routine incidence data alone (3–8).

The Sisyphus Study was designed to assess silent breast cancer prevalence in a consecutive forensic autopsy cohort in Lisbon, Portugal. Across the breast-focused pilot and parent publications, no imaging-detected silent breast cancers were identified in either the female or male cohorts (9–11). During the same autopsy programme, however, a number of histologically confirmed non-breast malignancies were recorded as unexpected secondary findings.

The present manuscript reports those incidental non-breast cancers as a stand-alone case series. The objective was to describe their frequency, anatomical distribution, and clinicopathological significance in the context of a consecutive forensic autopsy cohort.

## Methods

This study is a descriptive secondary analysis of incidental non-breast malignancies identified during the Sisyphus Study, a prospective forensic autopsy programme conducted at the National Institute of Legal Medicine and Forensic Sciences, Lisbon, Portugal, in collaboration with Hospital Sao Francisco Xavier (9–11).

The parent cohort included consecutive medicolegal autopsies performed between July 2016 and December 2019. Eligibility for the parent study has been described in detail elsewhere (9–11). For the present analysis, the key criteria were age 40 years or older, absence of major breast-region injury that would preclude parent-study sampling, and absence of known or clinically evident cancer at the time of case inclusion.

All decedents underwent full forensic autopsy. For the parent breast study, bilateral subcutaneous modified radical mastectomy was additionally performed in each case. Demographic variables and cause-of-death information were abstracted from the forensic file. All pathological findings identified during the autopsy, including unexpected extra-mammary tumours, were recorded systematically. Histological processing was undertaken in the pathology department of Hospital Sao Francisco Xavier.

For this report, an incidental cancer was defined as a histologically confirmed non-breast malignancy identified at autopsy in an individual without a prior clinical cancer diagnosis. Cases in which the previously unrecognised cancer had contributed directly to death were retained, because these represent clinically important silent disease rather than overdiagnosed indolent lesions.

Only descriptive statistics are reported. Prevalence was expressed as the proportion of incidental cancers within the male cohort, female cohort, and combined cohort.

## Results

The study cohort comprised 291 decedents: 74 males and 217 females. Mean age at death was 63.9 years in males and 65.5 years in females; mean body mass index was 28.63 kg/m^2^ and 24.89 kg/m^2^, respectively. The majority of decedents were of Caucasoid ethnicity (90.5% in males and 94.0% in females). No silent breast cancers were detected in the parent study cohort (9–11).

Fifteen incidental non-breast malignancies were identified overall, corresponding to a prevalence of 5.15% (15/291). Five malignancies were found among 74 male decedents (6.76%), and ten among 217 female decedents (4.61%) (Table 1).

**Table 1.**
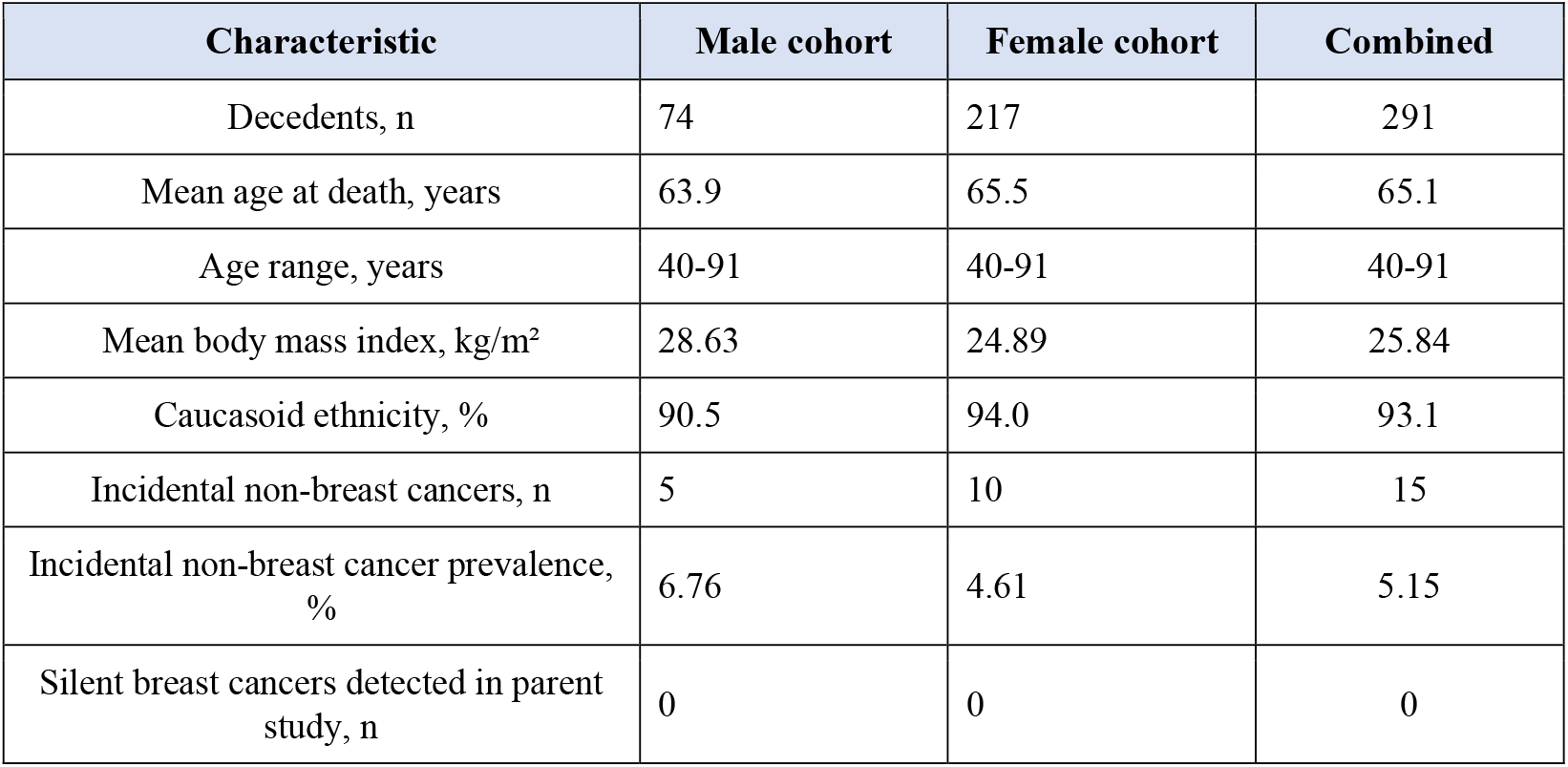
Cohort characteristics and principal study findings.

In the male cohort, the incidental tumours comprised one left colon adenocarcinoma, one right colon metastatic adenocarcinoma, one pancreatic metastatic adenocarcinoma, one gastric adenocarcinoma, and one splenic lymphoma. Thus, four of the five male cases involved the gastrointestinal tract, and two of the five were explicitly described as metastatic disease.

In the female cohort, the incidental tumours comprised three right colon adenocarcinomas, one perforated left colon adenocarcinoma, one left colon metastatic adenocarcinoma, one hepatic metastasis of left colon adenocarcinoma, two lung adenocarcinomas, one perforated gastric adenocarcinoma, and one ovarian metastatic adenocarcinoma. Colorectal malignancies therefore accounted for six of the ten female cases.

Across the combined cohort, colorectal cancer was the dominant tumour type, accounting for 8 of 15 malignancies (53.3%). At least five of the fifteen tumours were metastatic on autopsy diagnosis, and two gastrointestinal tumours were associated with perforation. These findings indicate that the silent cancer burden captured in this cohort included advanced disease rather than exclusively small or indolent lesions (Table 2).

**Table 2.** Incidental non-breast malignancies identified in the cohort.

| Sex | Primary site | Autopsy diagnosis | n |
| --- | --- | --- | --- |
| Male | Colorectal | Left colon adenocarcinoma | 1 |
| Male | Colorectal | Right colon metastatic adenocarcinoma | 1 |
| Male | Pancreatic | Pancreatic metastatic adenocarcinoma | 1 |
| Male | Gastric | Gastric adenocarcinoma | 1 |
| Male | Haematological | Splenic lymphoma | 1 |
| Female | Colorectal | Right colon adenocarcinoma | 3 |
| Female | Colorectal | Left colon adenocarcinoma (perforated) | 1 |
| Female | Colorectal | Left colon metastatic adenocarcinoma | 1 |
| Female | Colorectal | Hepatic metastasis of left colon adenocarcinoma | 1 |
| Female | Pulmonary | Lung adenocarcinoma | 2 |
| Female | Gastric | Gastric adenocarcinoma (perforated) | 1 |
| Female | Gynaecological | Ovarian metastatic adenocarcinoma | 1 |

## Discussion

This secondary analysis documents a measurable burden of clinically unrecognised non-breast malignancy in a consecutive forensic autopsy cohort. Fifteen histologically confirmed cancers were identified among 291 decedents, and the pathological spectrum included advanced colorectal, pancreatic, ovarian, pulmonary, gastric, and haematological disease. The findings are important because the parent cohort explicitly excluded individuals with known or clinically evident cancer, making these tumours invisible to routine clinical surveillance during life.

The broader autopsy literature supports the value of post-mortem material for understanding hidden disease reservoirs, but most relevant cancer-reservoir work has been site-specific rather than whole-body in scope (1–8). In that sense, the present series contributes additional evidence from a forensic setting in which decedents are not selected through oncology or tertiary-care pathways. A prior forensic autopsy study focused on tissue and organ safety reported a 7% incidence of unsuspected cancer in its practice, reinforcing the general principle that clinically occult malignancy is not rare in autopsy populations (2).

Colorectal cancer was the dominant incidental tumour type in the current series, representing 53.3% of all malignancies and 2.75% of the overall cohort. A recent systematic review of autopsy studies reported a pooled incidental colorectal cancer prevalence of 0.7% across heterogeneous populations, although direct comparison is limited by differences in age thresholds, sampling protocols, and case mix (12). Even with those caveats, the predominance of colorectal malignancy in the present cohort is clinically notable because colorectal cancer is one of the malignancies for which earlier detection can substantially reduce disease-specific mortality (13). Our findings are therefore consistent with missed opportunities for earlier detection, although individual screening histories were unavailable and no causal inference regarding screening participation can be made.

The two lung adenocarcinomas and the single ovarian metastatic adenocarcinoma also fit the wider literature on cancers that can remain clinically silent until late stages. A recent systematic review confirmed that subclinical lung cancer is regularly encountered at autopsy (14), while screening trials such as NELSON have shown that mortality reduction is achievable in selected high-risk populations rather than in the general population (15). By contrast, long-term follow-up from UKCTOCS did not demonstrate a statistically significant mortality benefit for general-population ovarian cancer screening (16), underlining why occult ovarian malignancy remains a persistent clinical challenge.

The gastric and pancreatic cases further strengthen the point that the hidden cancer burden is not limited to indolent lesions. In this cohort, gastrointestinal perforation and metastatic spread were already present in several cases. From a public-health perspective, forensic autopsy data may therefore complement, rather than compete with, registry-based surveillance by revealing the types of cancers that remain clinically missed until death.

Several limitations should be acknowledged. First, this was a single-centre secondary analysis derived from a parent study designed for breast-cancer ascertainment rather than systematic whole-body cancer mapping. Second, the absolute number of incidental cancers was small, precluding robust subtype comparisons. Third, Portuguese medicolegal constraints did not allow linkage to prior medical records, symptom history, or screening participation. Fourth, extra-mammary cancer ascertainment relied on standard forensic autopsy practice rather than protocolised organ-by-organ histological sampling, so the true prevalence of silent non-breast cancer may have been underestimated.

Despite these limitations, the present report provides a clinically meaningful description of unexpected extra-mammary cancers discovered in a prospective forensic autopsy cohort. It supports the argument that forensic autopsy findings deserve greater consideration within population cancer surveillance and cancer-control research.

## Conclusions

In summary, incidental non-breast malignancies were identified in 15 of 291 decedents in the Sisyphus forensic autopsy cohort, with colorectal cancer accounting for more than half of all cases. The series shows that clinically silent cancer in the general adult population can include metastatic and directly life-threatening disease. Larger multicentre autopsy collaborations with standardised pathological sampling and linkage to screening data would be valuable next steps.

## Data Availability

All data produced in the present study are available upon reasonable request to the authors

## Declarations

### Ethics approval

The parent Sisyphus Study was conducted under institutional and national medicolegal authorisations obtained in Portugal before tissue collection and data recording.

### Consent for publication

Not applicable.

### Data availability

Data are available from the corresponding author on reasonable request, subject to applicable legal and ethical restrictions.

